# Can tracking mobility be used as a public health tool against COVID-19 following the expiration of stay-at-home mandates?

**DOI:** 10.1101/2021.08.27.21262629

**Authors:** Peter Her, Sahar Saeed, Khai Hoan Tram, Sahir R Bhatnagar

## Abstract

**Background:** Considering the emergence of SARS-CoV-2 variants and low vaccine access and uptake, minimizing human interactions remains an effective strategy to mitigate the spread of SARS-CoV-2. The aim of this study was to create a novel multidimensional mobility index to capture the complexity of human interaction and evaluate its utility as an early indicator of surges in COVID-19 cases.

**Methods:** We used publicly available anonymous cell phone data compiled by SafeGraph, from all counties in Illinois, Ohio, Michigan and Indiana between January 1st to December 8, 2020. Six metrics of mobility were extracted for each county. Changes in mobility were defined as a time-updated 7-day rolling average. We used an unsupervised machine learning method known as functional principal component analysis (fPCA) to construct the latent mobility index (MI) using the six metrics of mobility. Associations between our MI and COVID-19 cases were estimated using a quasi-Poisson hierarchical generalized additive model adjusted for population density and the COVID-19 community vulnerability index.

**Results:** Individual mobility metrics varied significantly by counties and by calendar time. More than 50% of the variability in the data was explained by the first principal component by each state, indicating good dimension reduction. Following the expiration of stay-at-home orders, mobility increased across all counties and this was particularly evident on weekends. While an individual metric of mobility was not associated with surges of COVID-19, our MI was independently associated with COVID-19 cases in all four states given varying time-lags.

**Conclusion:** Following the expiration of stay-at-home orders, a single metric of mobility was not sensitive enough to capture the complexity of human interactions. Monitoring mobility can be an important public health tool, however, it should be modelled as a multidimensional construct.

## 1 Introduction

While highly effective vaccines are readily available in the United States, uptake remains low [1] and interventions aimed at minimizing human contact remain necessary to mitigate the spread of SARS-CoV-2 [2, 3, 4, 5]. Decreasing mobility patterns within populations, has been shown to be an effective strategy to curb infectious disease transmission. Reducing human interactions is particularly important given Delta (a variant of concern; that is highly transmissible) is the dominant SARS-CoV-2 strain driving the current pandemic wave in the United States.

The potential of monitoring population-level mobility patterns using geo-located mobile phone data as a public health tool has been demonstrated [6, 7, 8, 9]. In March 2020, worldwide adherence to lockdowns was measured using various mobility metrics [10, 11, 12, 13, 14, 15]. A modelling study from China showed 20-60% reductions in mobility notably controlled the spread of SARS-CoV-2 [16]. A study from Canada showed that reductions in mobility strongly predict future control of SARS-CoV-2 growth rates [5]. However, in the absence of social distancing interventions, the link between changes in population-level mobility and COVID-19 remains unclear [8, 9].

Population-level mobility, as it pertains to human interaction, is multidimensional. This is particularly true when assessing distinct geographical areas that vary by population density and socioeconomic factors across the United States [17, 18]. While the measurement of mobility is complex, studies to date have used single metrics such as the percentage of people remaining at home or changes in the distance travelled to summarize human interactions and evaluate trends and associations with COVID-19. These single metrics may oversimplify mobility associated with human interaction. As social distancing policies loosen from strict “lock down” to business-as-normal, the utility of continuously monitoring mobility will require a robust definition that is able to capture the complexity of population-level movement [19]. To this end, the aim of this study was to use advanced statistical methods to create a novel index that summarizes mobility as a latent construct using a combination of six mobility metrics. We evaluated how our mobility index varied across 365 counties in 4 states as a function of time. Finally, we assessed the performance of our mobility index by evaluating how mobility correlated with COVID-19 cases compared to a single metric from the time stay-at-home orders expired.

## 2 Methods

### 2.1 Data Sources

#### Mobility Metrics

We used aggregated mobility data publicly available through SafeGraph from January 1st to December 8, 2020, via the Social Distancing Metric database. SafeGraph uses a panel of GPS pings from anonymous mobile devices from a representative sample of the US Census population, to derive metrics of mobility. This data includes a range of spatial behaviors from >45 million mobile devices (≈ 10% of devices in the United States). To enhance privacy, SafeGraph excludes census block group information if fewer than two devices visited an establishment in a month.

*A priori*, we choose six mobility metrics commonly used in the literature as a proxy of human contact and that could be attributable to mobility behavior changes as associated with COVID-19 infections. Each metric is defined for a given day (t) for a given county (j). The metrics (s) included:

- The fraction of devices leaving home in a day
- The fraction of devices away from home for 3-6 hours (Part-time work behaviour)
- The fraction of devices away from home longer than 6 hours (Full-time work behaviour)
- The median time spent away from home
- The median distance traveled from home
- The average number of short stops (>3 stops for less than 20 min) (Delivery behaviours).

#### COVID-19 Cases

Confirmed COVID-19 cases data were retrieved from the New York Times open-source project [20]. This publicly available dataset aggregates county-level daily counts of diagnosed cases, from health services official reports.

#### Covariates

Demographic variables including population size and population density for each county was collected from the American Community Survey and the US Census Bureau. We used an aggregate measure of social and COVID-19 specific vulnerability to summarize socioeconomic status at the state and county-level, freely available as the COVID-19 community vulnerability index (CCVI) [21]. Dates for when stay at home orders were enforced and lifted were obtained from the State-level social distancing policies database [22]. There were several entries for each state due to policy revisions or updates. We used the first entry for each state in this database.

### 2.2 Analysis

#### Population

Given the magnitude of the available data, we reduced the number of states in our analysis by simply selecting the four most populous states in the Midwest. We used every county from each of the selected states to avoid any preferential selection.

#### Mobility Index

We defined mobility as a change of each mobility metric relative to the average of the week before (time-updated rolling average). For each county *j* = 1, …, 365, we index each of the 6 mobility metrics *s* = 1, …, 6 by calendar day *t* = 1, …, *m*_*j*_, where *m*_*j*_ is the total number of observed days since re-opening in county *j*. We define the following quantities:

- *X*_*j,t,s*_: the scalar value of mobility metric *s* measured on day *t* in county *j*.
- ***X***_*j,t*−8,…,*t−*1,*s*_: the value of mobility metric *s* measured on days *t* − 8, …, *t* − 1, i.e., the 7 days prior to day *t* in county *j*. This is a vector quantity.
- 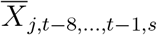: the average of the ***X***_*j,t*−8,…,*t*−1,*s*_.

The change from baseline mobility metric *s* for day *t* in county *j* is given by

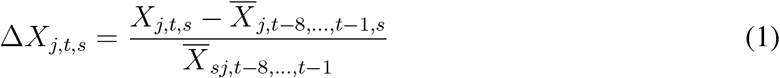

The use of a rolling average is unique to this analysis. Most studies have used a static relative baseline period such as mobility trends between January until February 2020 [6, 23, 24]. This common approach does not account for seasonal mobility variability or changes as a result of the pandemic [25, 26, 27]. In contrast our baseline (rolling average) takes into consideration temporal trends that were likely changing with evolving public health policies.

Since our hypothesis was each metric could be attributed to a common underlying notion of mobility, we used an unsupervised machine learning method known as functional principal component analysis (fPCA) to create our latent mobility index [28]. Briefly, PCA is a technique for reducing the dimensionality of multiple variables while minimizing information loss. This is done by creating new uncorrelated variables (principal components) that successively maximize variance. A “functional” PCA accounts for the longitudinal nature of the data. We applied fPCA on Δ*X*_*j,t,s*_ separately for each county and extracted the first principal component, i.e., the linear combination of individual mobility metrics that explained the most variance. We denote this first principal component by fPCA_*j,t*_, a score summarizing mobility in each county (*j*) on a given day (*t*). To enable comparability between counties and states, fPCA_*j,t*_ was scaled as Z-scores, which defined our mobility index (MI) given by:

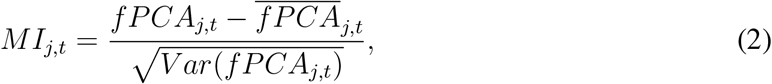

where 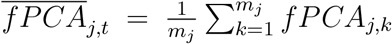 and *V ar*(*f PCA*_*j,t*_) are the average and variance of the fPCA scores in county *j* over the observed time period, respectively.

The interpretation of *MI* is as follows, *MI* = 0, on average there was no change in mobility relative the previous week; *MI* = 1 on average there was an increase in mobility by one standard deviation relative to the last week, and *MI* = −1 on average there was a decrease in mobility by one standard deviation relative to the last week. An animation was created to visualize the relative daily changes of *MI* by counties.

#### Association with COVID-19

For each county *j*, let *y*_*j,t*_ be the number of confirmed COVID-19 cases on day *t* = 1, …, *m*_*j*_, and **q**_*j,t*_ = [*MI*_*j,t*−0_, …, *MI*_*j,t*−21_] denote the vector of lagged occurrences of our mobility index (defined in Equation (2)) with 0 days and 21 days as minimum and maximum lags, respectively. In words, the first element of **q**_*j,t*_ represents the value of our mobility index on day *t*, the second element represents the value of our mobility index one day prior to *t*, and so on. From the time “stay at home” orders expired until December 8th 2020, the relationship between daily counts of COVID-19 cases (*y*_*j,t*_) and mobility (**q**_*j,t*_), accounting for up to 21 days of lag, was estimated with a quasi-Poisson hierarchical generalized additive model (HGAM) [29, 30] of the form:

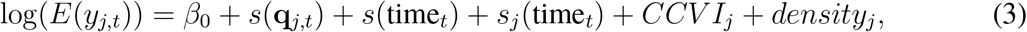

where *β*_0_ is the intercept, *s*(·) are the smooth non-parametric functions of the predictor variables, *CCV I*_*j*_ is the COVID-19 community vulnerability index and *density*_*j*_ is the population density (people per square kilometer) at the county-level. The term *s*(**q**_*j,t*_) in Equation (3) captures the potentially non-linear and delayed effect of mobility on COVID-19 cases through a cross-basis function [31]. We used penalized cubic regression splines [29] for both dimensions, with interior knots placed at Z-scores of −3, −2, −1,0,1,2,3 for *MI*_*j,t*_, and 7 and 14 days for the lag. Given the heterogeneity of COVID-19 epidemiology across counties, models included both a state level calendar time effect *s*(time_*t*_) using thin plate regression splines [32] and a county level calendar time effect *s*_*j*_(time_*t*_) using a factor-smoother interaction basis [30]. Population size was used as offset in each model. A separate model was run for each of the selected states.

There were two main advantages for using a HGAM to evaluate the association between mobility and COVID-19 cases: (1) it can quantify the non-linear functional relationships over time where the shape of each function varies across counties, and (2) it has the capacity of modelling varying lags [33]. For example, it is estimated that it takes a median of 5 days from SARS-CoV-2 infection until the onset of symptoms, followed by an unknown number of days before people get tested and a positive infection is confirmed. This lag can be differential at both the patient-level (develop symptoms and get tested) and also at the county-level (lag in reporting tests). Given this variation, we were able to control for varying lagged exposures (up to 21 days) at the county-level. To evaluate the utility of our mobility index, we compared a dose response relationship between mobility and COVID-19 cases and goodness of fit statistics of our latent MI compared to a single measure of mobility (the fractions of devices leaving the home). All analyses were performed using R version 4.0.2 [34] along with the mgcv [29] and dlnm [35] packages. Code and data for reproducing all the results, figures and animation in this paper is available at https://github.com/sahirbhatnagar/covid19-mobility.

## 3 Results

### 3.1 Mobility patterns

Daily mobility changes of three hundred sixty-five counties from the four most populous states in the Midwest: Illinois, Ohio, Michigan and Indiana were analyzed between January 1 2020 until December 8 2020. State-level sociodemographic and economic characteristics were similar across four states and are summarized in Table 1.

**Table 1:** The sociodemographic and economic characteristics of Illinois (IL), Ohio (OH), Michigan (MI) and Indiana (IN).

| State | Order | Lift | Population | Number of counties | Median household income | Cumulative cases per capita at opening<sup>a</sup> | Cumulative cases per capita until Dec 8<sup>th</sup> <sup>a</sup> |
| --- | --- | --- | --- | --- | --- | --- | --- |
| Illinois | March 21 | April 8 | 12,741,080 | 102 | \$65,030 | 118 | 6324 |
| Ohio | March 24 | April 7 | 11,689,442 | 88 | \$56,111 | 41 | 4363 |
| Michigan | March 24 | April 14 | 9,995,915 | 83 | \$56,697 | 269 | 4427 |
| Indiana | March 25 | April 7 | 6,691,878 | 92 | \$55,746 | 83 | 5909 |

<sup>a</sup> cumulative cases per 100,000 population

Figure 1 illustrates the average daily changes of the six mobility metrics between January and December 2020 of each state (average of all counties) relative to the week before. Overall, each metric had a unique trajectory but trends were similar across four states. Based on the average change, the number of devices not at home and delivery behavior (more than 3 stops lasting for less than 20 mins) remained stable throughout time. While changes in work-related metrics and the median time devices remained were not at home varied more. Of the four states, mobility changes were more pronounced in Ohio. Across all states, relative to the previous seven days, mobility increased daily between March and May. Mobility metrics varied considerably by counties (S1, S2, S3, S4), illustrating how aggregating changes at the state-level may mask granular changes at the county-level.

**Figure 1:**
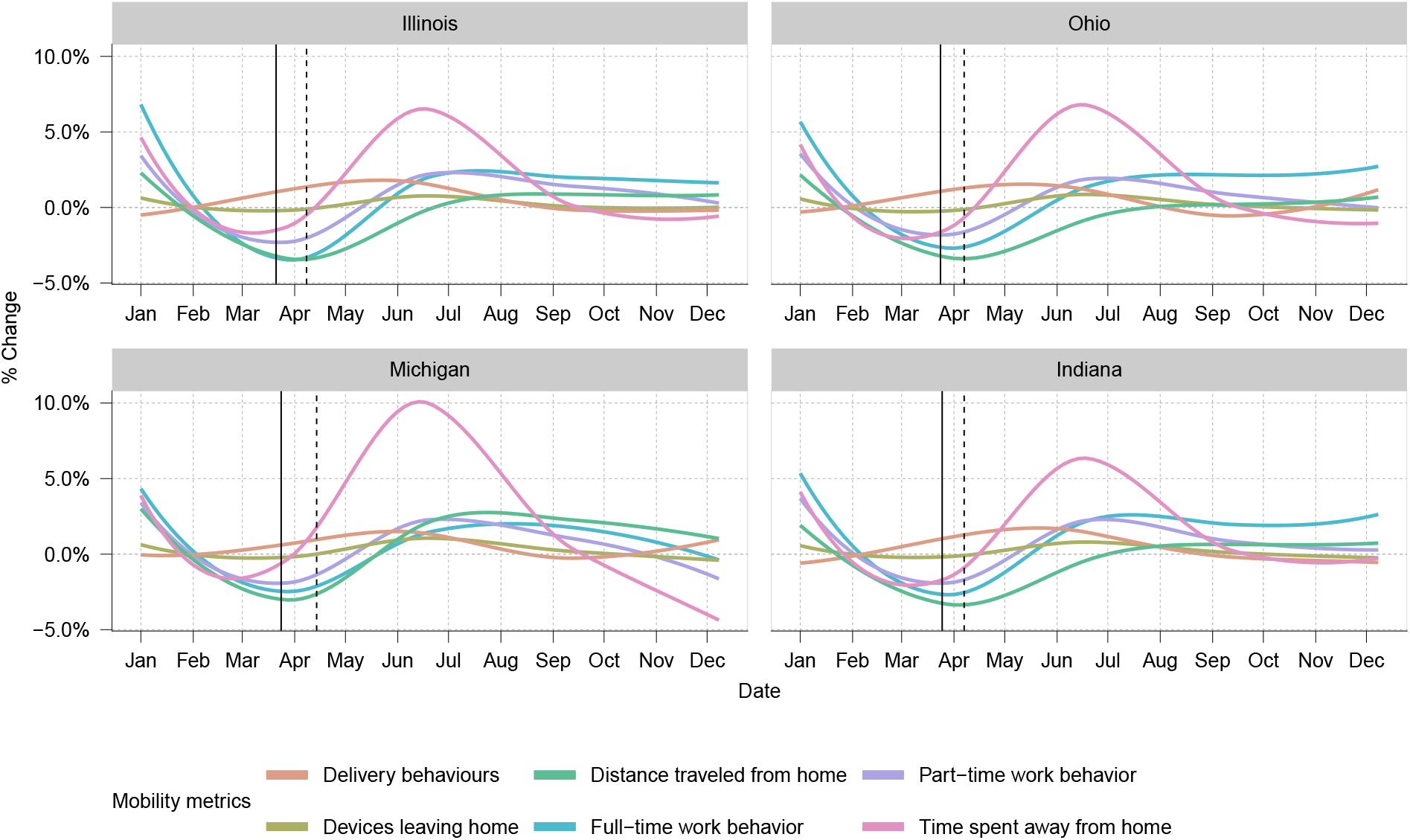
The average daily changes from baseline in the six mobility metrics for all counties of each state between January and December 2020. The baseline was calculated using a rolling average of the 7 previous days. The solid vertical lines represent the date the stay-at-home orders were put in place while the dotted vertical lines represent the dates the stay-at-home orders were lifted.

### 3.2 First fPCA summarizes mobility patterns by counties

We created a latent index of mobility by counties as given by Equation (2) which is derived from the first fPCA. Table 2 provides the median and inter quartile range of the proportion of variance explained by the first fPCA across counties in a given state. We see that over 50% of the variance is explained by the first fPCA for a majority of all the counties analyzed, indicating good dimension reduction. In Supplemental Figures S5, S6, S7 and S8, we provide the absolute correlations between our MI and each individual metric by county for Illinois, Ohio, Michigan and Indiana, respectively. We see the correlations are particularly strong with full/part time work behaviour as well as time spent away from home. Importantly, there was significant heterogeneity across counties which would otherwise be missed when aggregating mobility metrics at the state level.

**Table 2:** Median (inter quartile range) of the proportion of variance explained by the first fPCA by state. *n* represents the number of counties in each state.

| Illinois ($n = 102$) | Ohio ($n = 88$) | Michigan ($n = 83$) | Indiana ($n = 92$) |
| --- | --- | --- | --- |
| 0.57 (0.50, 0.63) | 0.71 (0.67, 0.74) | 0.61 (0.53, 0.69) | 0.65 (0.59, 0.70) |

Figure 2 compares the changes of the MI from the day stay at home policies expired and July 4^*th*^ (Independence Day). Blue shades indicate MI <0 (decrease in mobility) and red shade indicate MI>0 (increase in mobility). This graph provide some evidence that our MI is appropriately capturing mobility as we would expect there to be more movement on a traditionally busy U.S. holiday compared to earlier on in the pandemic when stay at home orders were lifted. In the Supplemental material, we also provide an animation illustrating the daily changes from reopening to December 8^*th*^. The animation shows substantial difference in mobility patterns across counties that vary from day to day. The most dramatic change over time is increases in mobility from a weekday to a weekend.

**Figure 2:**
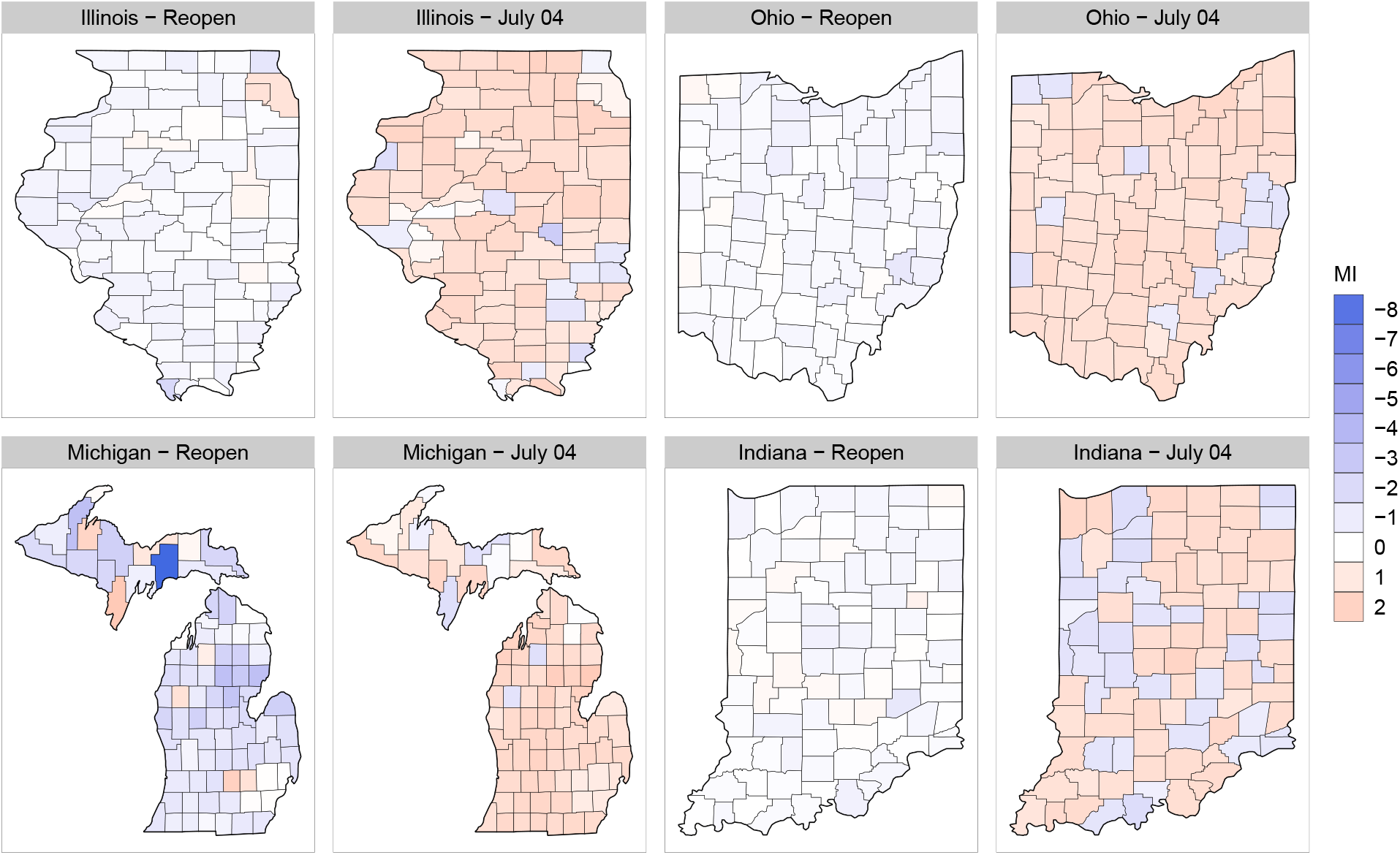
MI values for each county of each state on the day the stay-at-home orders expired (reopen) and on July 4^*th*^, 2020. Blue shades indicate a decrease in mobility (MI < 0) and red shades indicate an increase in mobility (MI > 0).

### 3.3 Association with COVID-19

To evaluate the utility of the MI, we compared its association with COVID-19 cases and a commonly used single metric of mobility (fraction of devices leaving home) (Figure 3). Notably, the single metric was not associated with COVID-19 cases in any state at any lagged time point. While all four states showed a clear dose response of MI and COVID-19 cases following a 10-21 day lag. Across all four states the MI model resulted in significantly better goodness of fit statistics compared to the single metric (Table 3).

**Table 3:** Analysis of deviance table comparing the goodness of fit between the MI model (fPCA) and the fraction of devices leaving home (single) of the four states. Degrees of freedom shown is for the *χ*^2^ test statistic.

| State | Residual deviance | Degrees of freedom | Reduction in deviance for the MI model (fPCA) |
| --- | --- | --- | --- |
| Illinois (single) | 152,581 |  |  |
| Illinois (fPCA) | 149,282 | 3.6 | 3,299 |
| Ohio (single) | 123,551 |  |  |
| Ohio (fPCA) | 112,975 | 8.6 | 10,576 |
| Michigan (single) | 261,759 |  |  |
| Michigan (fPCA) | 250,888 | 7.0 | 10,871 |
| Indiana (single) | 83,517 |  |  |
| Indiana (fPCA) | 80,297 | 1.7 | 3,220 |

**Figure 3:**
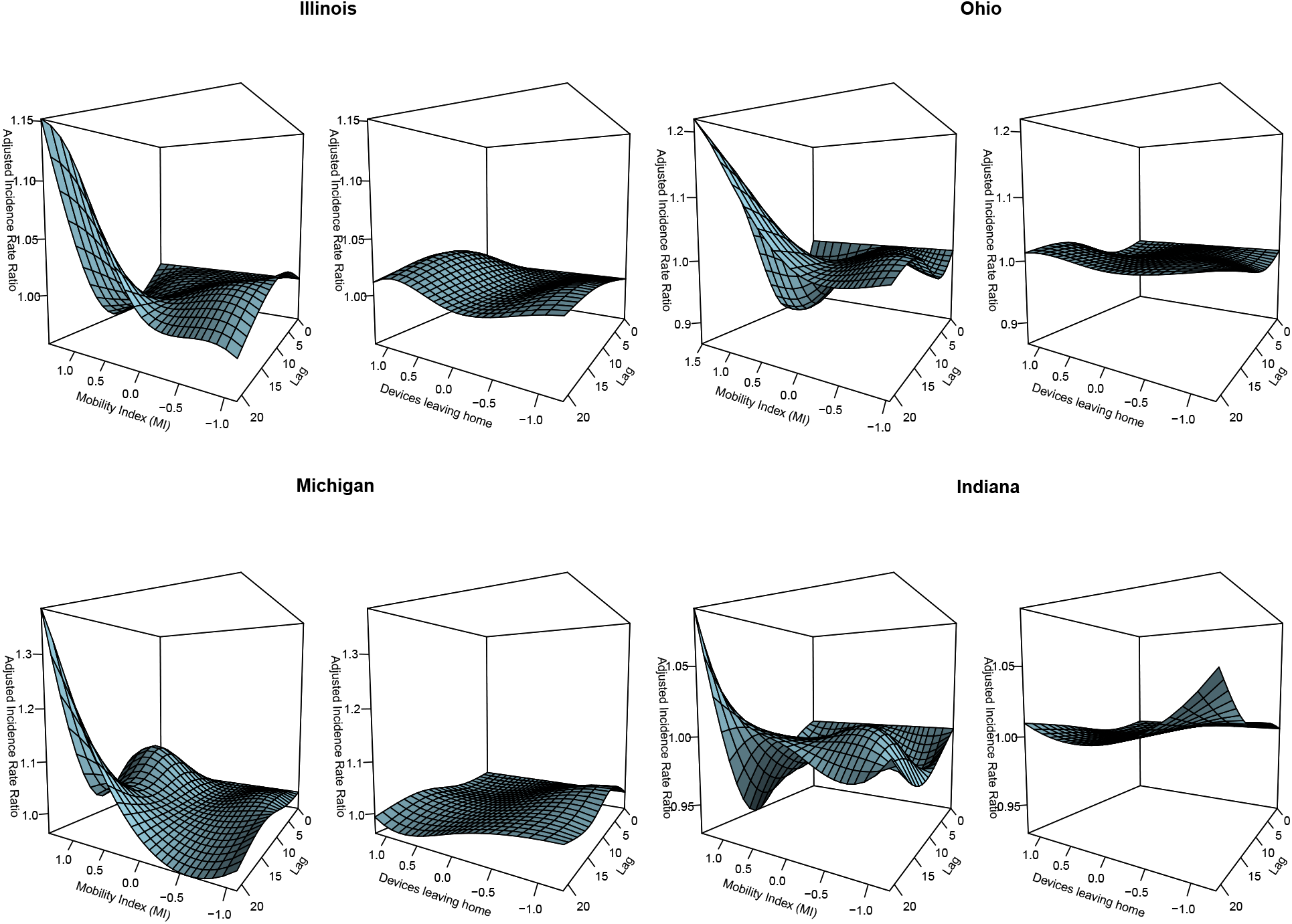
Model results comparing the MI and its association with COVID-19 cases and a commonly used single metric of mobility (fraction of devices leaving home). For each state, the left panel summarizes the multidimensional MI; the right panel represents the percentage of devices leaving their home (x-axis); y-axis is the adjusted incidence rate ratio of COVID-19, at varying lagged response (0-21 days) (z-axis).

## 4 Discussion

The COVID-19 pandemic is now fueled by highly transmissible variants of concern. Understanding the association between mobility and disease transmission can help tailor non-pharmaceutical interventions to mitigate outbreaks and potentially be used as an early indicator for surges in new infections. We leveraged freely available cell phone data with an unsupervised machine learning approach to create a multidimensional index of mobility. Results from our study suggest following the expiration of stay-at-home physical distancing policies, single metrics of mobility were not sensitive enough to capture the complexity of human mobility related to disease transmission. Our MI was correlated with COVID-19 cases for Illinois, Ohio, Michigan and Indiana. In comparison, the single metric of mobility (fraction of devices leaving home) was not associated with incident cases. Our results also demonstrate the importance of evaluating changes at a granular level as there was significant heterogeneity within states.

The use of mobility data was suggested to be a powerful tool to determine the impact of public health policies [19]. During the initial phase of the pandemic, several studies examined the association between mobility and COVID-19 [2, 36, 37, 38]. For example Lasry et al. [6] found an association between changes in mobility (% personal mobile devices leaving home) at the state-level and COVID-19 cases during the first COVID-19 wave. However at this time NPI were more homogenous across counties and states. In contrast our study was the first to evaluate the association of mobility and COVID-19 cases following the expiration of stay-at-home orders reflecting mobility behavior that is more reflective of typical population-level movement. We demonstrated among hundreds of counties from four states, time-updated relative changes were associated with increases in COVID-19 cases. Furthermore, results from our study suggest our mobility index should be considered an important confounder when evaluating other non-pharmaceutical interventions.

The strength of our study was the use multiple advanced statistical methods to measure mobility and evaluate its association with COVID-19 cases. The fPCA used to create the mobility index effectively captured the heterogeneity of the individual metrics over time and across counties within a given state. The unsupervised nature of this approach prevented the model from overfitting when evaluating the association with cases. Furthermore, we modelled a non-linear functional relationship between mobility and COVID-19 cases using a HGAM model while simultaneously fitting different lagged time periods. The expectation that the lag time should vary across states was confirmed by our results. The use of these methods has been under appreciated in the epidemiological and public health studies; we provide code and data to expand the use as we believe these methods could have wide applications in future research.

Our study also has limitations. Although cell phone data was freely available and could help to predict trends during the pandemic, it is only a proxy for human contact. In this study we attempted to define a more robust definition of mobility, however it still remains a surrogate exposure. The association between mobility and COVID-19 cases may be underestimated, given our outcome is dependent on testing. Testing capacity has significantly changed throughout the pandemic in the United States. Seroprevalence studies estimate case detection is underrepresented by a factor of three times [39]. Although we do not believe this underrepresentation to be differential, outcomes such as COVID-19 related deaths and hospitalizations may be less bias. While the advantage is clear, the utility of these outcomes as a “real-time” public health tool is debatable as the latency period (time of infection to outcome) is long (>21 days). As with all observational studies, associations should not be interpreted causally. Our model does not take into consideration confounding interventions that could also increase or mitigate transmission such as the proportion of the population adhering to physical distancing guidelines, wearing masks, interactions outside vs inside or air quality. To effectively measure social distancing patterns individual wearable technology or trackers, would be more sensitive compared to aggregate data, but this raises ethical and privacy concerns [40]. Recent reports have hypothesized the COVID-19 pandemic may not be following a normal distribution but over dispersed or driven by “super spreader” transmission events which we did not account for in our model [41].

## 5 Conclusion

Our study underscores the potential of using freely available cell phone data as public health tool. We show changes in mobility can be used a predictor of surges in COVID-19 cases. However monitoring mobility in the absence of strict non-pharmaceutical interventions such as “stay at home” will require robust definitions.

## Data Availability

Code and data for reproducing all the results, figures and animation in this paper is available at https://github.com/sahirbhatnagar/covid19-mobility

## A County-level mobility metrics

**Figure S1:**
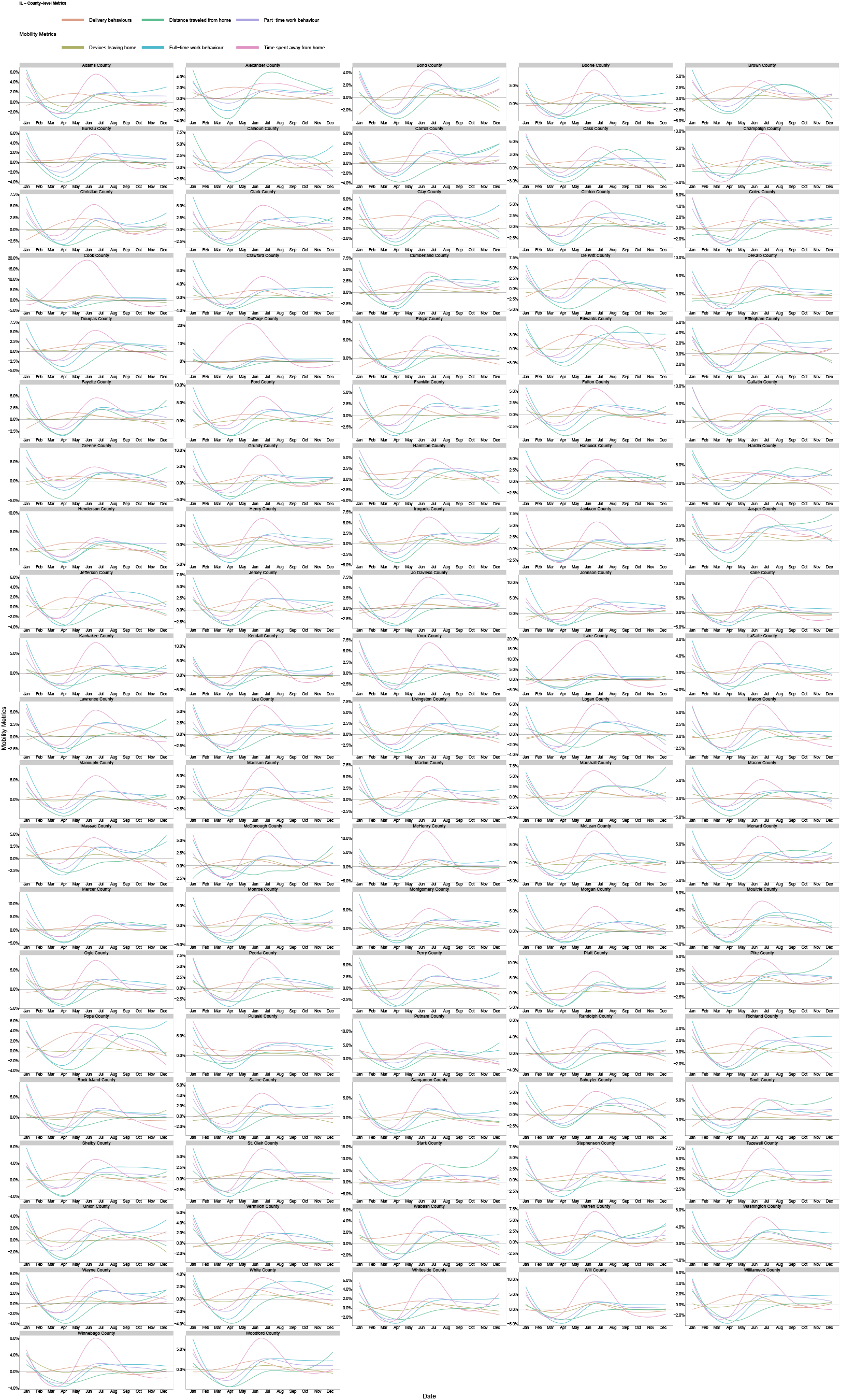
Illinois

**Figure S2:**
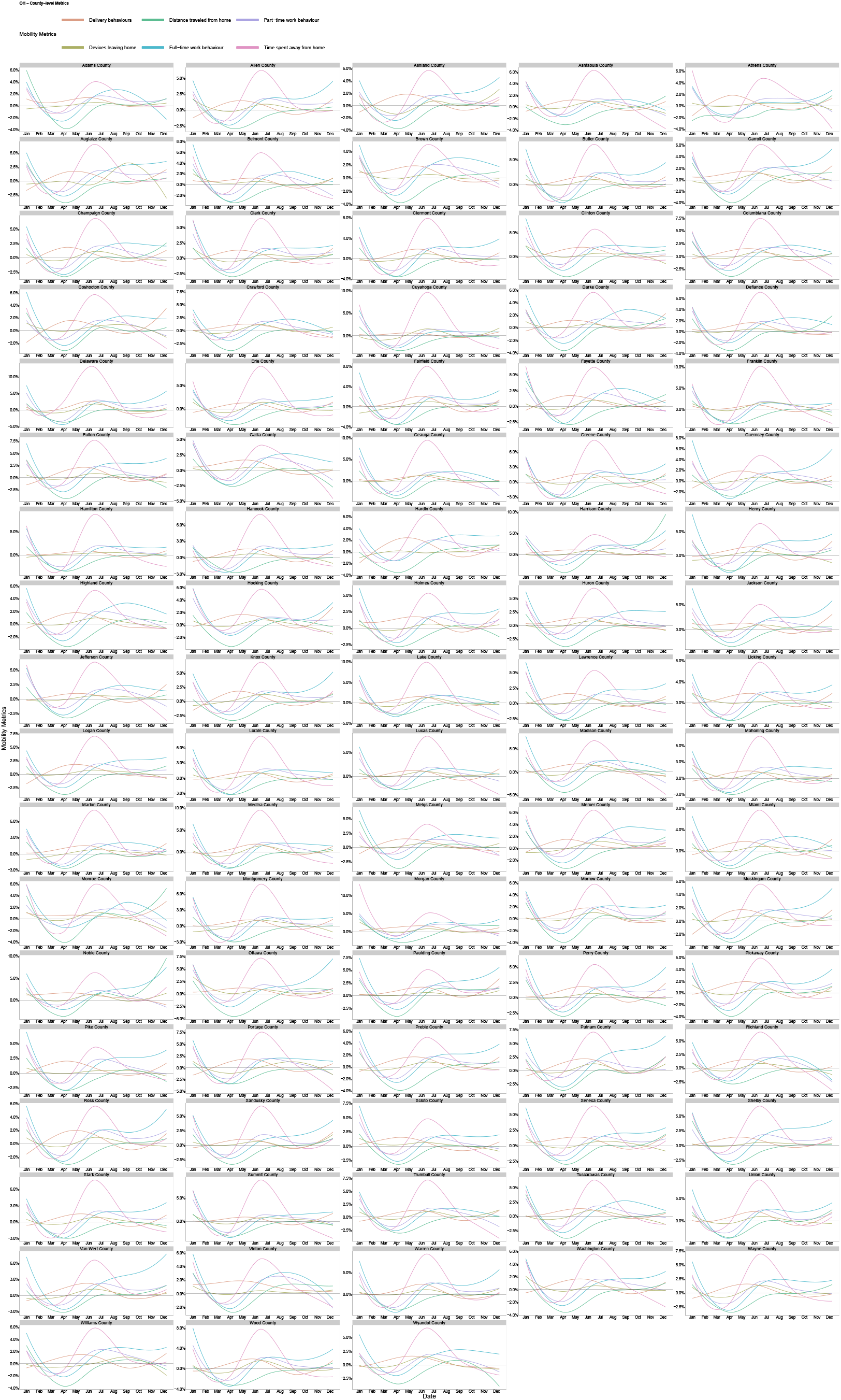
Ohio

**Figure S3:**
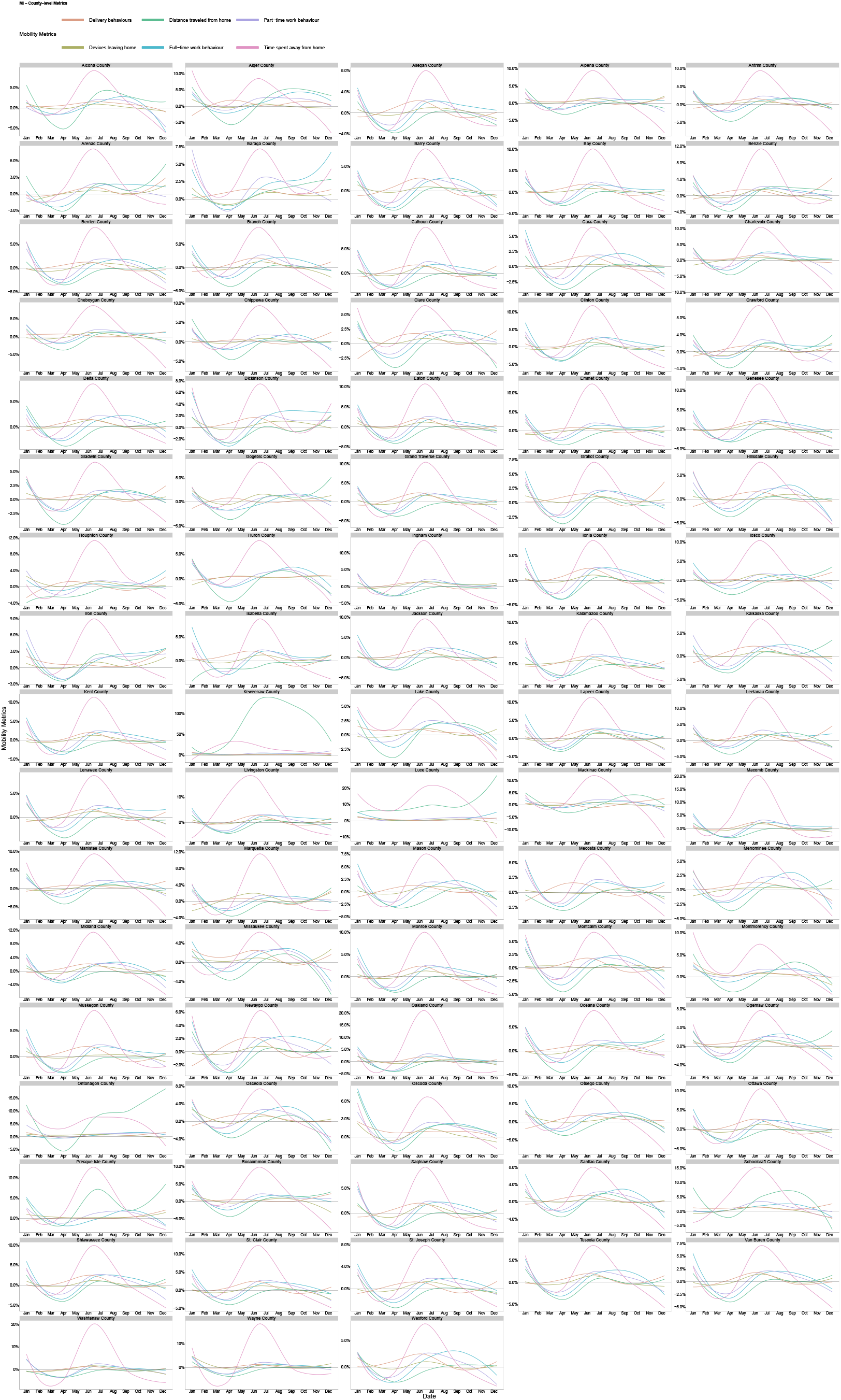
Michigan

**Figure S4:**
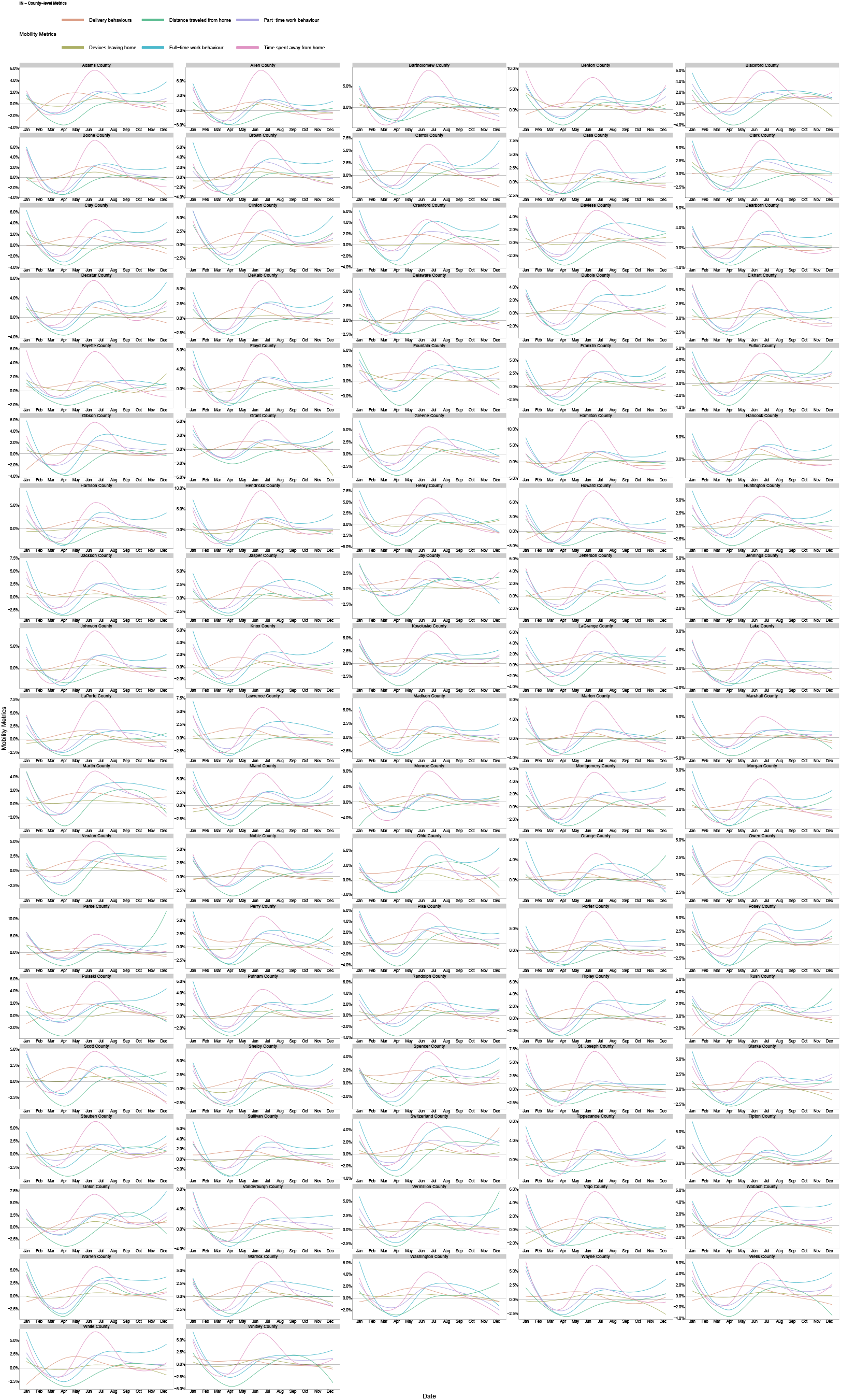
Indiana

## B Correlations between MI and individual metrics

**Figure S5:**
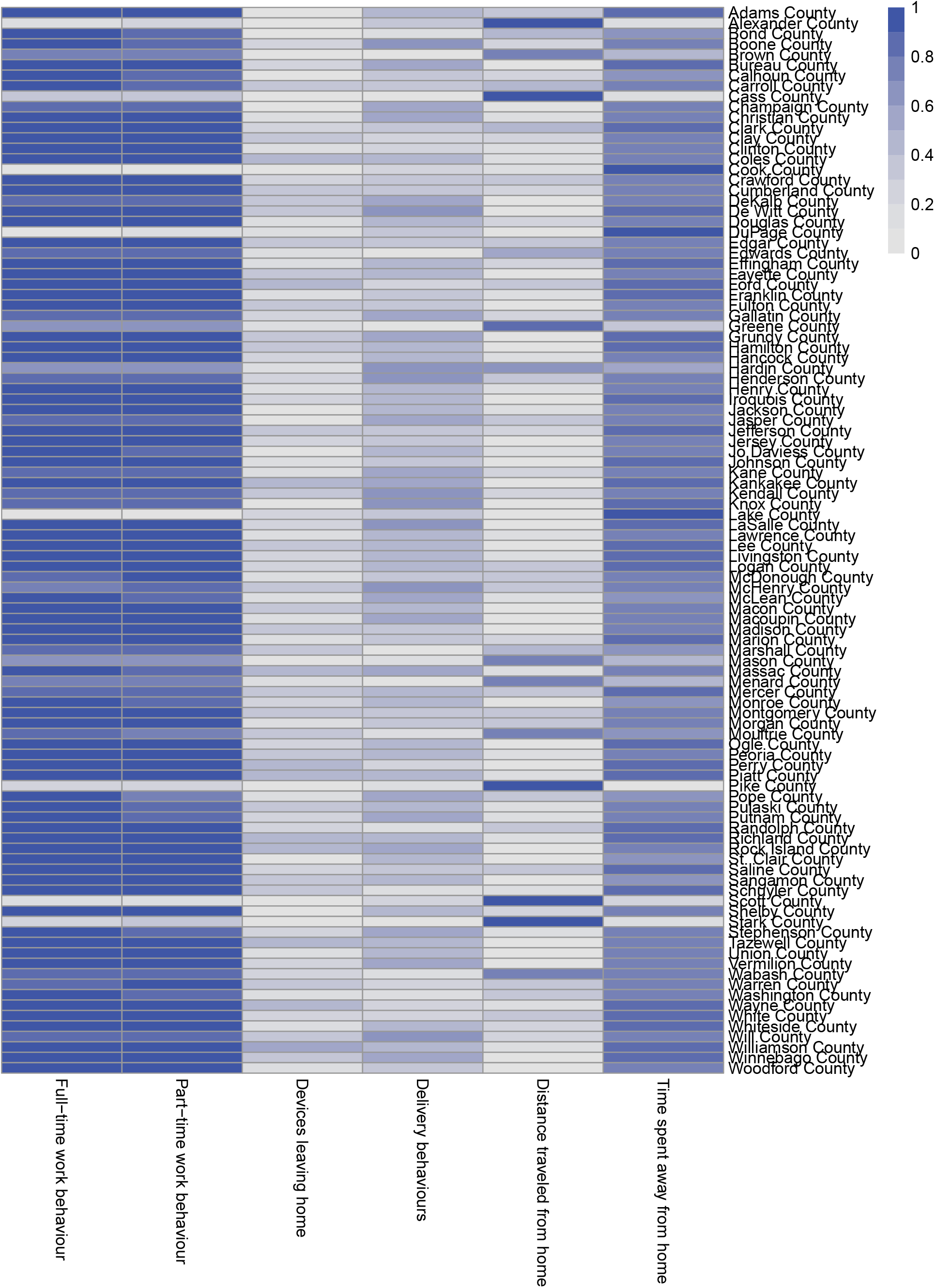
Illinois

**Figure S6:**
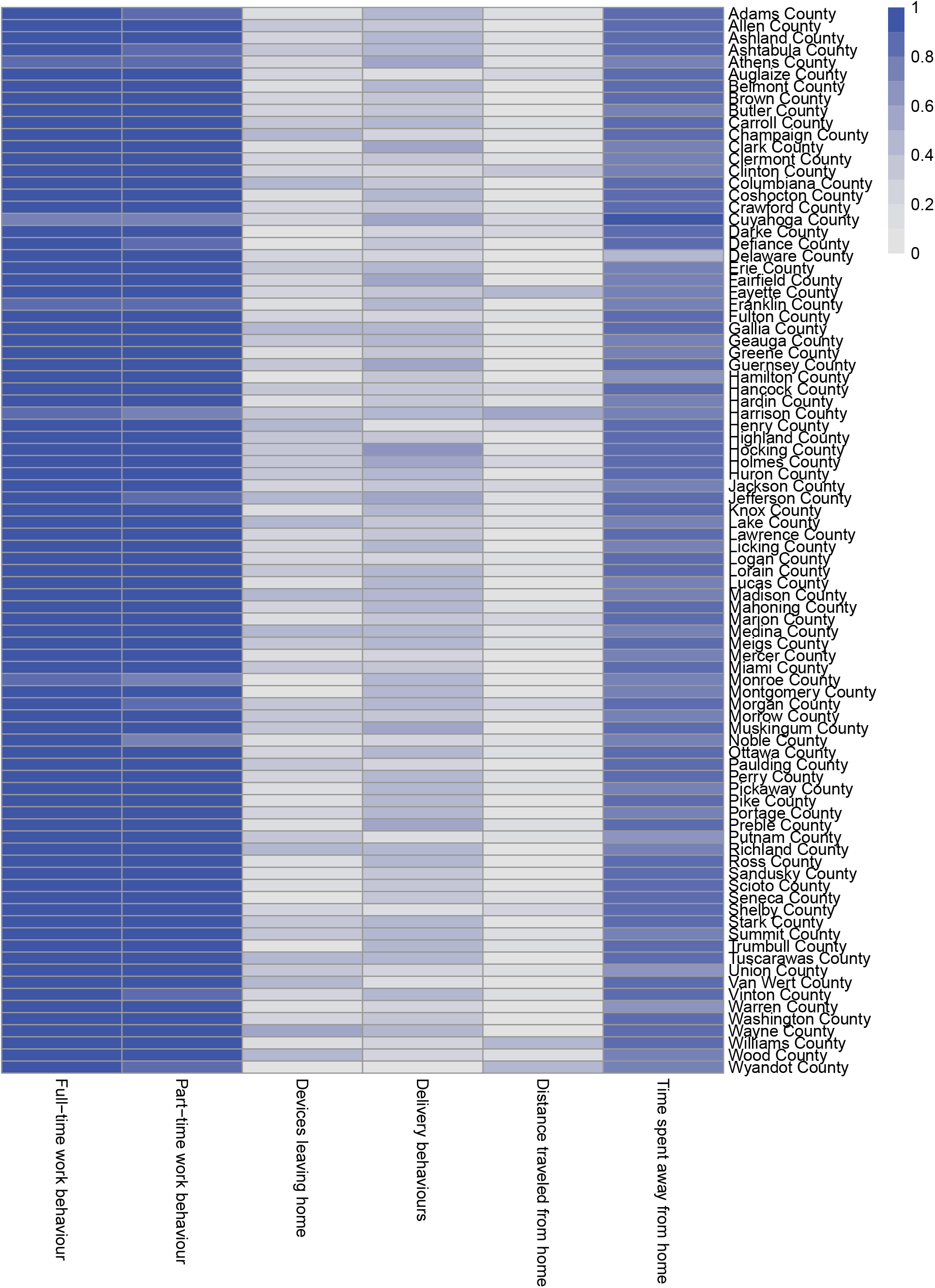
Ohio

**Figure S7:**
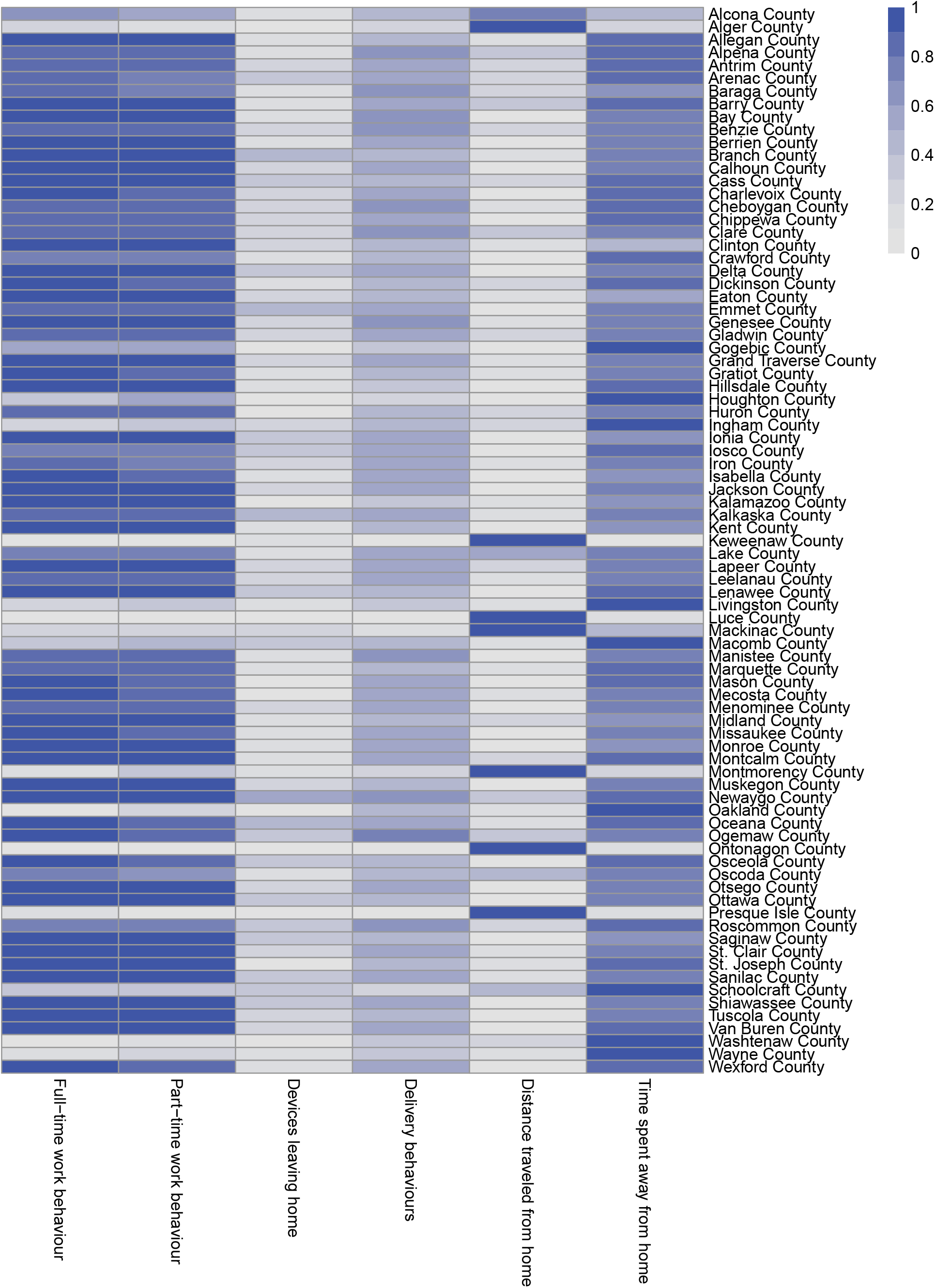
Michigan

**Figure S8:**
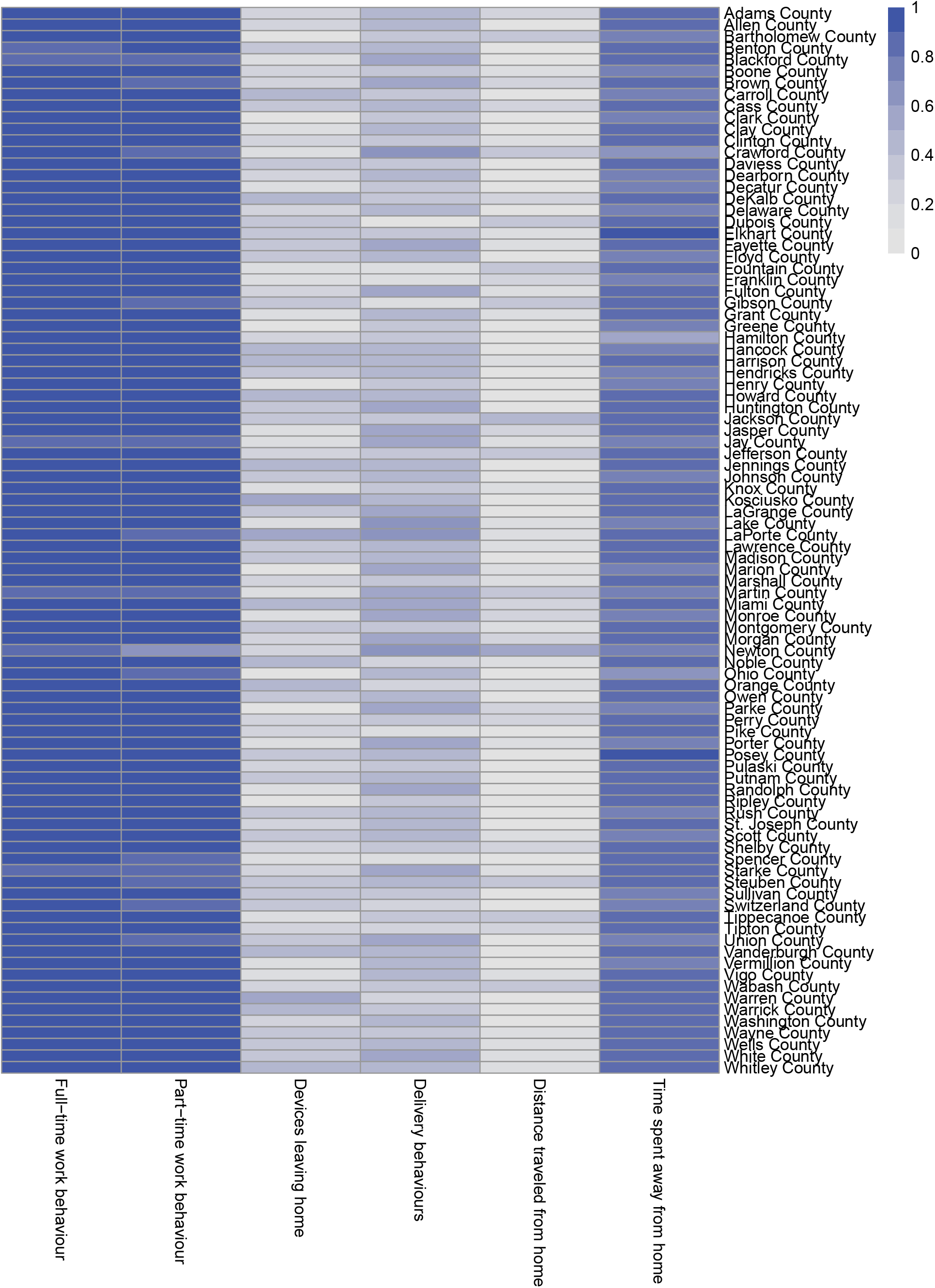
Indiana

